# Treatment outcomes of bedaquiline-resistant tuberculosis: a retrospective and matched cohort study

**DOI:** 10.1101/2025.01.21.25320876

**Authors:** Lindokuhle Mdlenyani, Zahraa Mohamed, Jacob AM Stadler, Nomfuneko Mtwa, Graeme Meintjes, Robin Warren, Johanna Kuhlin, Sean Wasserman

**Affiliations:** Department of Health, Eastern Cape Province, South Africa; Centre for Infectious Diseases Research in Africa, Institute of Infectious Disease and Molecular Medicine, University of Cape Town, South Africa; Department of Medicine, University of Cape Town, South Africa; Blizard Institute, Queen Mary University of London, London, United Kingdom; Division of Molecular Biology and Human Genetics, SAMRC Centre for Tuberculosis Research, Faculty of Medicine and Health Sciences, Stellenbosch University, Cape Town, South Africa; Department of Medicine Solna, Karolinska Institutet, Stockholm, Sweden; Department of Infectious Diseases, Karolinska University Hospital, Stockholm, Sweden; Institute for Infection and Immunity, City St George’s, University of London, London, United Kingdom

## Abstract

**Background:** Rising prevalence of bedaquiline resistance undermines benefits from this life-saving drug for rifampicin-resistant tuberculosis (RR-TB). Despite increasing awareness, patient-level outcomes for bedaquiline-resistant TB have not been well-characterised and case management poorly defined.

**Methods:** We conducted a retrospective cohort study with matched controls at a TB referral hospital in South Africa. Cases included patients ≥13 years with a phenotypic bedaquiline-resistant *Mycobacterium tuberculosis* isolate identified between January 2018 and June 2023. Matched controls (1:1) with bedaquiline-susceptible RR-TB were selected from a prospective observational study conducted at the same facility. Primary outcomes included time to sputum culture conversion (SCC), a modified WHO-defined unfavourable outcome, and TB-free survival (alive, with SCC, and in care or treatment completed) through 18 months. Adjusted analyses used Cox proportional hazards models.

**Findings:** Eighty-two patients with bedaquiline-resistant TB were included; 57 (70%) were HIV-positive and 17 (21%) had no prior exposure to bedaquiline or clofazimine. Bedaquiline was prescribed for 72 (88%) and meropenem (plus amoxicillin-clavulanate) for 32 (39%) after submission of the sputum sample with the first bedaquiline-resistant *Mycobacterium tuberculosis* isolate. At 18 months, 43 (52%) patients achieved TB-free survival; 17 (20%) failed to achieve sustained SCC, there were 19 (23%) deaths, and 50 (80%) survivors (n=63) were still on treatment. WHO treatment outcomes following treatment initiation were unfavourable in 54 (67%) patients, driven by treatment failure in 35 (43%). Median time to SCC after treatment initiation was 175 (IQR 100–254) days in the bedaquiline resistant cohort and 32 days (IQR 30–42 days) in the matched bedaquiline susceptible cohort. Baseline smear microscopy grade (HR 0·41, 95% CI 0·23–0·73, p=0·003) and bedaquiline resistance (HR 0·06, 95% CI 0·02–0·23, p<0·001) were associated with reduced SCC.

**Interpretation:** Current treatment options for bedaquiline-resistant TB result in prolonged therapy, delayed microbiological responses, and poor clinical outcomes.

**Funding:** This work was partially supported by the South African Medical Research Council (grant number MRC-RFA-SHIP 02–2018). JK was funded by Swedish Heart & Lung Foundation (20220859 and 20240774) Swedish Society of Medicine (SLS-985976). SW was supported by the National Institutes of Health (U01AI170426) and the Bill & Melinda Gates Foundation. GM was supported by the Wellcome Trust (214321/Z/18/Z, and 203135/Z/16/Z), and the South African Research Chairs Initiative of the Department of Science and Technology and National Research Foundation (NRF) of South Africa (Grant No 64787).

## Background

Bedaquiline has been transformational in improving outcomes and enabling treatment shortening for rifampicin-resistant tuberculosis (RR-TB). ^1–4^ In 2023, an estimated 176000 people were treated for RR-TB, most of whom received bedaquiline as recommended by the World Health Organization (WHO) and almost 60 national TB programmes (NTPs) had implemented the 6-month bedaquiline-linezolid-pretomanid-moxifloxacin (BPaL/M) regimen. ^1^ The South African NTP has been providing bedaquiline to most people with RR-TB since 2018 after its incorporation into a standardised 9–12-month oral regimen. ^5^ Current national guidelines recommend BPaL/L (which includes levofloxacin instead of moxifloxacin), now provided to most patients with RR-TB. ^6^

Expanded use has been accompanied by emergence of bedaquiline resistance, posing a threat to the progress made over the past decade. ^1^ Resistance-associated genetic variants causing phenotypic resistance, emerge during and after treatment, driven by sub-therapeutic bedaquiline exposure due to its long elimination half-life. ^7,8^ These resistance mutants can be transmitted in communities. ^9^ Bedaquiline resistance may worsen treatment outcomes, shown both in routine care^4^ and among patients from clinical trials evaluating BPaL, ^10^ potentially undermining global effectiveness of shorter oral treatment for RR-TB.

Prevalence of bedaquiline resistance appears to be increasing. South African surveillance data from 2015–2019 (n=2023) showed 3·8% pre-treatment phenotypic bedaquiline resistance that was strongly associated with prior bedaquiline exposure: 21% versus 3·6% resistance in those exposed compared treatment-naïve patients. ^4^ In a more recent survey of *Mycobacterium tuberculosis (M. tuberculosis)* isolates from unselected patients with RR-TB (March to November 2023), the South African National Institute for Communicable Diseases found 3·6% to 10·2% bedaquiline resistance in three provinces in South Africa. ^11^ A study conducted in Mozambique (n=704) reported prevalence of genotypic bedaquiline resistance increasing from 3% in 2016 to 14% in 2021. ^12^

Optimal treatment of bedaquiline resistant TB is not known. The WHO and the South African NTP recommend an individualised regimen guided by extended drug susceptibility testing (DST) and, in South Africa, consultation with a national clinical advisory committee. ^6,13,14^ These so-called ‘salvage regimens’ usually rely on the addition of WHO group C drugs including parenteral carbapenems. ^15^

Reliance on bedaquiline-based treatment for RR-TB, coupled with increasing population prevalence of bedaquiline resistance and limited evidence to guide clinical management, requires improved understanding of the clinical impact from bedaquiline resistance. This retrospective and matched cohort study aims to describe the management strategies and treatment outcomes of patients with bedaquiline-resistant pulmonary TB in a high HIV-burden setting in South Africa.

## Methods

### Study design and population

We conducted a retrospective cohort study of patients with bedaquiline-resistant RR-TB, matched to controls with RR-TB and confirmed bedaquiline susceptibility. All patients were treated within the South African NTP at Nkqubela Chest Hospital (NCH), a regional referral centre for drug-resistant TB in the Eastern Cape province, South Africa. Until March 2023 the South African NTP performed bedaquiline phenotypic DST (pDST) for selected RR-TB cases, including those with poor treatment responses or known resistance to fluoroquinolones, second-line injectable drugs, or isoniazid; after March 2023 all *M. tuberculosis* isolates from RR-TB cases were tested. ^4,11^

The retrospective cohort included patients ≥13 years at NCH diagnosed with sputum culture positive pulmonary TB and found to have a bedaquiline-resistant *M. tuberculosis* isolate on pDST between January 2018 and June 2023. The date range was selected to identify cases after established bedaquiline use in the NTP and to capture 18-month follow-up information. Individuals without available treatment information at NCH were excluded.

A control group with RR-TB and confirmed bedaquiline susceptibility was selected from a prospective observational study (SHIFT-TB, n=260) that evaluated programmatic outcomes with an oral bedaquiline-based 9–12-month regimen at the same facility and received treatment within an overlapping time period (January 2021 to August 2022). ^16^ Controls were matched 1:1 with bedaquiline resistant cases on age, HIV status, and baseline culture status (see Supplementary methods). No participants enrolled in the SHIFT-TB study had prior RR-TB treatment as this was an exclusion criterion for the regimen being evaluated.

In the SHIFT-TB cohort, written informed consent or assent and parental consent for participants <18 years was sought before participation in the study. Since we used routinely collected data for the bedaquiline-resistant cohort, a waiver for consent and assent was approved by ethics committees.

### Procedures

We reviewed hospital files, pharmacy records, the National Health Laboratory Service (NHLS) database, and EDRWeb for patients in the bedaquiline-resistant cohort. For matched controls, we extracted data from the SHIFT-TB database. Information included demographics, comorbidities, microbiological data, previous TB treatment, antituberculosis medication, treatment outcomes, and vital status until 18 months follow-up time or study end (20 January 2025). Data were entered into a bespoke Research Electronic Data Capture database (REDCap®) (Vanderbilt University, Nashville, TN, USA).

All microbiological samples for *M. tuberculosis* were processed by the NHLS, including routine testing with rapid molecular tests and culture using Mycobacterial Growth Indicator Tube (MGIT, Becton and Dickson, Franklin Lakes, NJ, USA). ^17^ Bedaquiline pDST was conducted using MGIT at the NHLS and at the Division of Molecular Biology and Human Genetics, Stellenbosch University. A critical concentration of 1mg/L was used^18^ with quality control carried out including *M. tuberculosis* H37Rv daily (NHLS) or on each new test batch (Stellenbosch University).

### Outcomes and definitions

We analysed three treatment outcomes: time to sputum culture conversion (SCC), a modified WHO-defined unfavourable outcome, and TB-free survival.

SCC was defined as the date of the first of two sputum samples with negative *M. tuberculosis* cultures, consecutive or not, without any intervening positive culture. Reversion included any single positive culture following SCC. Our definition of SCC was less stringent than the 2021 WHO definition, considering less frequent sputum collection when using data from routine care in the bedaquiline-resistant cohort. ^19^

We measured time to SCC from ‘treatment initiation’, defined relative to the date of collection of the first bedaquiline-resistant *M. tuberculosis* isolate (referred to as the ‘index sputum’). For patients receiving treatment prior to the index sputum, treatment initiation date was reassigned to the date of the index sputum. For patients not yet on treatment at the index sputum, the treatment start date was recorded as the date of treatment initiation. Time to *first SCC* was defined as the time to the first occurrence of conversion, irrespective of subsequent reversion. Time to *sustained SCC*, considered SCC only if it was maintained without reversion. This distinction accounted for patients with fluctuating culture status, with the *sustained SCC* reflecting stable conversion.

Unfavourable treatment outcome was adapted from the WHO 2021 definition (see Supplementary Methods for details) ^13^. As there is no defined treatment duration for bedaquiline-resistant TB, we used a patient-specific endpoint of treatment cessation for at least two months for outcome reporting.

TB-free survival was defined as a composite of sustained SCC, being alive, and either having completed treatment or being in care for TB.

### Statistical analysis

Demographic and clinical characteristics were described using summary statistics. Medication duration was assessed over the full period of treatment or until administrative censoring at study end (20 January 2025). Incomplete data were handled using a complete case approach (cases without complete data were excluded from respective analyses).

For the bedaquiline-resistant cohort, we estimated proportions with TB free survival at 6, 12, and 18 months; time to first and sustained SCC over 12 months after treatment initiation; time to death, censored at 18 months; and modified WHO-defined treatment outcomes. TB free survival and time to death were assessed both from the date of collection of the index sputum and from the date of treatment initiation to account for patients for whom treatment initiation was delayed. As a measure of treatment response, time to SCC and WHO treatment outcomes were assessed from treatment initiation only.

Cox proportional hazards were estimated for time to sustained SCC and mortality, and the following potential confounders were evaluated for adjustment: age, sex, body mass index (BMI), HIV and antiretroviral (ART) status, CD4 count, baseline microscopy positivity, and previous treatment for RR-TB. Bedaquiline duration, from the date of index sputum, was included as a continuous variable in the mortality analysis and as a binary variable in the SCC analysis due to bidirectional causation (i.e., those with a longer time to SCC were more likely to receive bedaquiline for a longer period because this outcome informed treatment decisions). For variables of interest that did not meet the proportional hazards assumption, we estimated Kaplan-Meier survival curves stratified by the relevant variable. Multivariable Cox proportional hazards models for SCC and mortality included variables that were significantly associated with the outcome on univariable analysis (p<0·05). To mitigate immortal time bias, a sensitivity analysis for mortality considered only those patients who survived at least eight weeks after the index sputum.

Matched participants from SHIFT-TB were pooled with the bedaquiline-resistant cohort and a stratified Cox regression model was used to account for matching. Cox proportional hazards were estimated for time to sustained SCC with adjustments considered for sex, BMI, baseline microscopy grade, baseline fluoroquinolone resistance, and bedaquiline resistance; this was followed by multivariable analysis including significant predictors (p<0·05). Previous RR-TB treatment could not be assessed as a confounder due to collinearity since none of the controls had previously been treated for RR-TB. Time to SCC was quantified by restricted mean survival time over 12 months from treatment start. Kaplan-Meier estimates for time to death, stratified by bedaquiline resistance, were compared with the log rank test.

All statistical analysis was performed using RStudio Version 2024·09·0+375 (Posit team, Boston, MA, USA).

### Ethics

Ethical approval was obtained from the Human Research Ethics Committee, University of Cape Town (REF. 887/2023 and 690/2019) and the Eastern Cape Department of Health Ethics Committee (EC_202311_022 and EC_201911_017).

## Results

We identified 86 patients with bedaquiline-resistant pulmonary TB, 82 of whom were included in the analysis (Table 1). Reasons for exclusion were missing hospital records (n=2) and treatment at a different institution (n=2).

**Table 1:** Demographic and clinical characteristics of patients with bedaquiline-resistant tuberculosis at the time of index sputum collection^1^.

| Characteristic | N = 82<sup>2</sup> |
| --- | --- |
| Age, years | 38 (30, 46) (14, 74) |
| Female sex | 42 (51%) |
| Weight, kg | 52 (44, 61) (33, 96) |
| BMI, kg/m<sup>2</sup>, n = 81 | 18.4 (16.8, 21.7) (13.4, 35.4) |
| Sputum microscopy positive, n = 81 | 45 (56%) |
| Baseline microscopy grade, n = 81 |  |
| 0 | 36 (44%) |
| 1+ | 20 (25%) |
| 2+ | 14 (17%) |
| 3+ | 11 (14%) |
| HIV test positive | 57 (70%) |
| ART status (if HIV positive) |  |
| Currently on ART | 51 (89%) |
| Not on ART | 6 (11%) |
| HIV viral load<sup>3</sup> (if HIV positive) |  |
| Below level of detection | 13 (33%) |
| Detectable | 27 (68%) |
| Quantitative viral load<sup>3</sup>, n=27 | 6600 (1200, 117600) (100, 3520000) |
| CD4 count<sup>3</sup>, cells/mm<sup>3</sup>, n=40 | 167 (82, 335) (19, 1170) |
| Previous RR-TB episodes |  |
| 0 | 42 (51%) |
| ≥ 1 | 40 (49%) |
| Previous RR-TB outcome lost to follow up | 15 (18%) |
| Previous RR-TB outcome treatment failure | 22 (27%) |
| Previous treatment with bedaquiline or clofazimine | 65 (79%) |
| Previous treatment with bedaquiline | 64 (78%) |
| Previous treatment with clofazimine | 65 (79%) |
| Number of initial antituberculosis drugs when starting treatment after the collection of the index sputum | 6 (5, 7) (3, 8) |

<sup>1</sup>Index sputum is defined as the first bedaquiline-resistant *Mycobacterium tuberculosis* isolate
<sup>2</sup>Median (Q1, Q3) (Min, Max); n (%)
<sup>3</sup>Within six months before and after the time of collection of the bedaquiline-resistant sputum
ART = Antiretroviral treatment, BMI = body mass index, RR-TB = Rifampicin-resistant tuberculosis

At the time of index sputum, 71 (87%) patients fulfilled criteria for extensively drug-resistant TB (XDR-TB; resistance to both bedaquiline and fluoroquinolones), ^13^ two of whom had *M. tuberculosis* isolates that were additionally resistant to linezolid (Supplement Table 1). Of the *M. tuberculosis* isolates tested at some point after the index sputum, 7/33 (21%) had linezolid resistance. Phenotypic clofazimine resistance was present in 67 (92%) of the baseline isolates. 19 (23%) patients had a bedaquiline-susceptible result prior to the index sputum date. The cohort included six patients from two sets of families (three members each) with similar resistance profiles within families.

The treatment regimen following the index sputum included bedaquiline for 72 (88%) patients, provided for a median duration of 5·5 months (range: 4 days–13·5 months) (Table 2). Over 90% were prescribed clofazimine, linezolid and terizidone. Meropenem (plus amoxicillin-clavulanate) was included in the regimen for 32 (39%) patients, started at a median 156 (IQR 115 to 236) days after the index sputum and prescribed for a median duration of 5·6 (IQR 4·4–6·5) months. Median overall treatment duration following the index sputum was 17·5 (IQR 10·4–20·4) months.

**Table 2:** Treatment regimens and treatment duration of the index episode during which bedaquiline resistance was identified.

| Characteristic | Before index sputum<sup>1</sup><br>N = 45<sup>2</sup> | After index sputum<sup>1</sup><br>N = 82<sup>2</sup> |
| --- | --- | --- |
| Bedaquiline | 43 (96%) | 72 (88%) |
| Clofazimine | 45 (100%) | 79 (96%) |
| Linezolid | 42 (93%) | 79 (96%) |
| Terizidone | 36 (80%) | 78 (95%) |
| Delamanid | 18 (40%) | 61 (74%) |
| Para-aminosalicylic acid | 18 (40%) | 61 (74%) |
| Levofloxacin | 35 (78%) | 53 (65%) |
| Pyrazinamide | 28 (62%) | 43 (52%) |
| Meropenem (plus amoxicillin-clavulanate) | 3 (6.7%) | 32 (39%) |
| Isoniazid | 18 (40%) | 27 (33%) |
| Ethambutol | 21 (47%) | 25 (30%) |
| Moxifloxacin | 3 (6.7%) | 14 (17%) |
| Ethionamide | 5 (11%) | 6 (7.3%) |
| Rifabutin | 2 (4.4%) | 5 (6.1%) |
| Time to meropenem initiation after index sputum collection, days | NA | 156 (115, 236) (1, 576) |
| Time to meropenem initiation after treatment start, days | NA | 140 (45, 213) (0, 576) |
| Duration of meropenem treatment, days | 32 (4, 181) (4, 181) | 171 (135, 199) (32, 457) |
| Duration of linezolid treatment, days | 82 (16, 153) (1, 574) | 391 (173, 583) (4, 858) |
| Time of bedaquiline treatment, days | 120 (30, 169) (1, 380) | 167 (94, 215) (4, 413) |
| Treatment duration, days | 121 (31, 251) (1, 819) | 539 (317, 620) (4, 1379) |
| Treatment modification<sup>3</sup> | NA | 44 (54%) |

<sup>1</sup>Index sputum is defined as the first bedaquiline-resistant *Mycobacterium tuberculosis* isolate
<sup>2</sup>n (%); Median (Q1, Q3) (Min, Max)
<sup>3</sup>Defined as the addition of at least two new drugs after the bedaquiline-resistant sputum collected
NA = Not applicable

Median time to first SCC was 100 days (95% CI 89–179) and time to sustained SCC was 175 days (95% CI 100–254) (Supplement Figure 1, Figure 1). None of the variables tested (age, sex, BMI, HIV status, HIV combined with ART status, previous RR-TB, baseline microscopy positivity, or bedaquiline use) were significantly associated with SCC (Supplement Table 2). Therefore, a multivariable analysis was not conducted.

**Figure 1:**
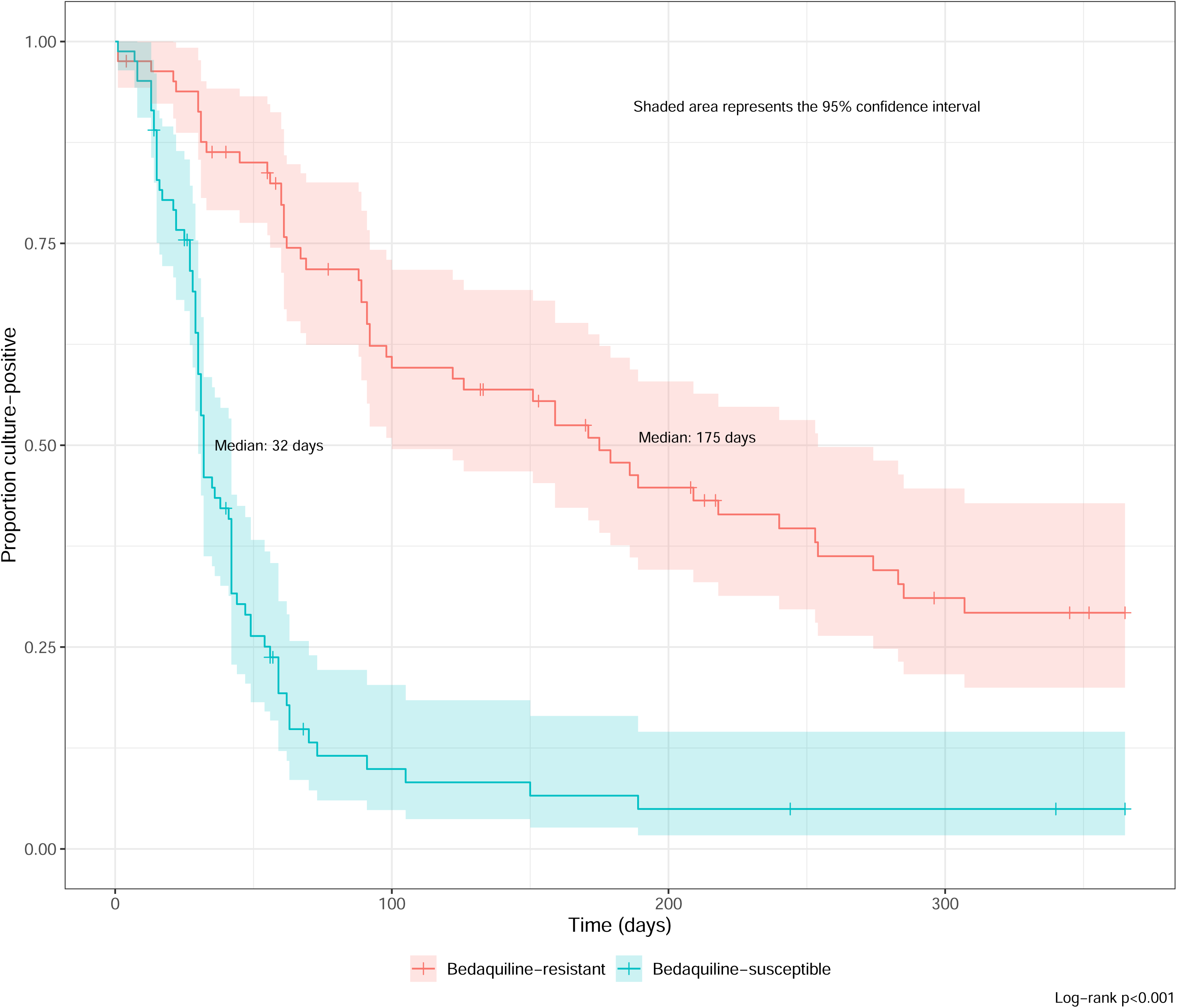
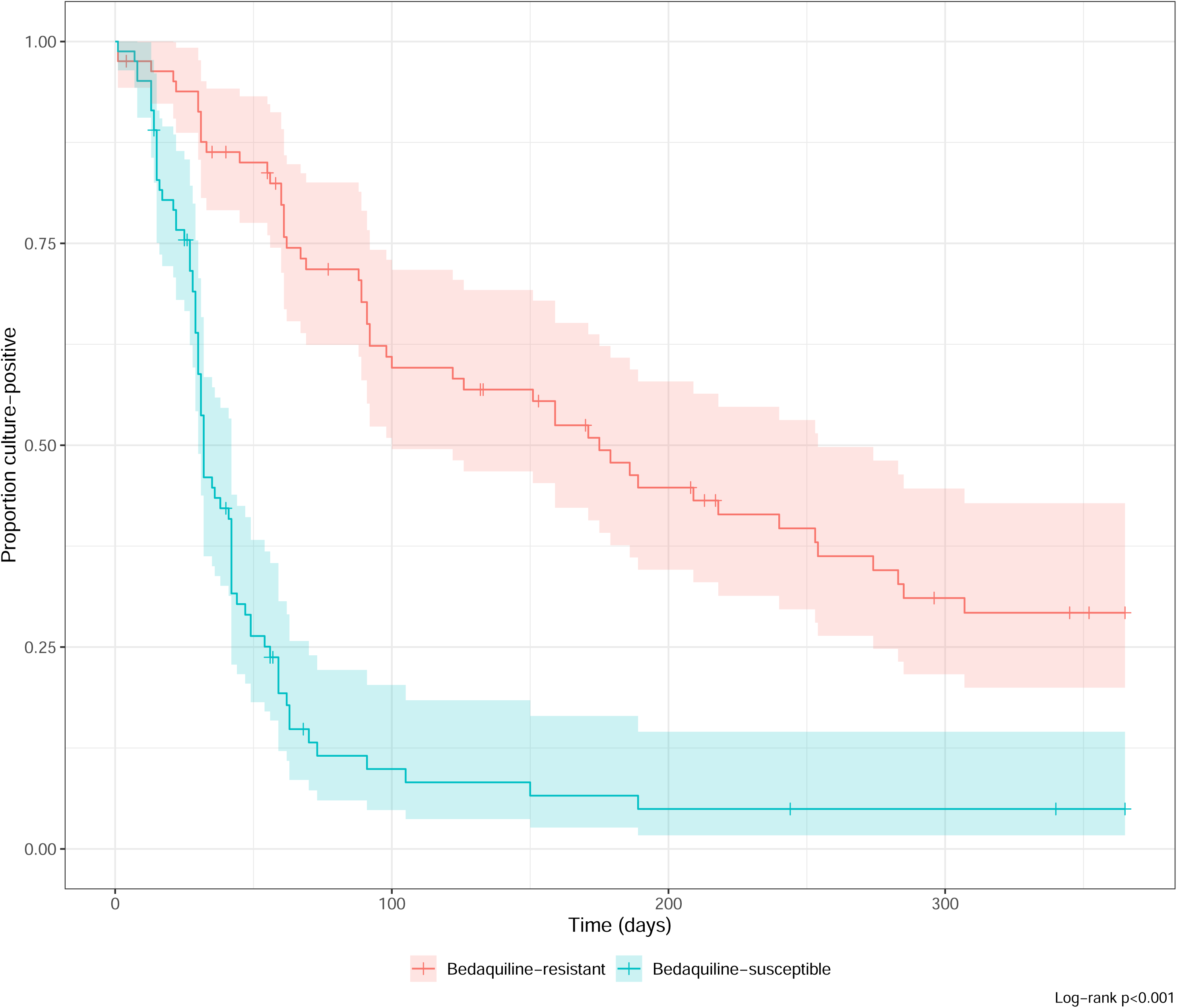
Kaplan–Meier curves for sustained sputum culture conversion after treatment initiation in the combined cohort of bedaquiline–resistant cases and matched bedaquiline–susceptible controls, by bedaquiline resistance and censored at 12 months.

TB-free survival after the index sputum was achieved for 41 (50%), 47 (57%) and 43 (52%) patients at 6, 12, and 18 months, respectively (Table 3). No patients had completed treatment at 6 months and 50 (80%) of survivors (n=63) were still on treatment at 18 months. At 6 months, 8 (10%) patients had died, increasing to 19 (23%) at 18 months. 43 (52%) and 46 (56%) patients were alive with sustained SCC at 6 months and 18 months, respectively. Results were similar when assessed from treatment initiation following the index sputum (Supplement Table 3).

**Table 3:** Tuberculosis-free survival at 6, 12, and 18 months after index sputum^1^ collection in patients with bedaquiline-resistant tuberculosis.

| Characteristic | Month 6 N =<br>82<sup>2</sup> | Month 12 N =<br>82<sup>2</sup> | Month 18 N =<br>82<sup>2</sup> |
| --- | --- | --- | --- |
| Achieved | 41 (50%) | 47 (57%) | 43 (52%) |
| Alive, TB free & treatment complete<sup>3</sup> | 0 (0%) | 2 (2.4%) | 8 (9.8%) |
| Alive, TB free & in care (treatment ongoing) | 41 (50%) | 45 (55%) | 35 (43%) |
| Not achieved | 41 (50%) | 35 (43%) | 39 (48%) |
| Not alive (i.e. died) | 8 (9.8%) | 13 (16%) | 19 (23%) |
| NOT TB free, alive & in care (treatment ongoing) | 26 (32%) | 18 (22%) | 15 (18%) |
| NOT in care<sup>4</sup> | 7 (8.5%) | 4 (4.9%) | 5 (6.1%) |
| Alive, TB free | 2 (2.4%) | 1 (1.2%) | 3 (3.7%) |
| Alive, NOT TB free | 5 (6.1%) | 3 (3.7%) | 2 (2.4%) |

<sup>1</sup>Index sputum is defined as the first bedaquiline-resistant *Mycobacterium tuberculosis* isolate
<sup>2</sup>n (%)
<sup>3</sup>TB-free is defined as achieving sputum culture conversion without culture reversion by the specified timepoint.
<sup>4</sup>Lost to follow up (irrespective of TB status) or completed treatment but not TB-free
TB = Tuberculosis

Only longer duration of bedaquiline use (HR 0·75, 95% CI 0·63– 0·90 per month; p-value (p) 0·002) was associated with reduced mortality, therefore, a multivariable analysis was not conducted (Supplement Table 4). In the sensitivity analysis including patients who survived at least eight weeks after collection of the index sputum, both BMI (HR 0·84, 95% CI 0·71–0·99 per unit increase; p =0·040) and bedaquiline duration use (HR 0·82, 95% CI 0·69–0·99 per month; p=0·039) were associated with reduced mortality; the effect size was maintained when combined in a multivariable model, but the association lost statistical significance (Supplement Table 5). Use of meropenem was not associated with survival (Supplement Figure 2).

Treatment outcome according to the modified WHO definition measured at the end of treatment was unfavourable in 54 (67%) patients (Supplement Table 6). Treatment failure, driven by lack of culture conversion by 6 months, accounted for unfavourable outcomes in 35 (43%) patients and 8 (10%) were lost to follow-up.

Baseline characteristics were similar between the bedaquiline-resistant and bedaquiline-susceptible cohorts, except higher baseline microscopy positivity and more advanced HIV disease (median CD4 count 100 cells/mm^3^) among people with HIV in the bedaquiline-susceptible cohort (Supplement Table 7).

Median time to sustained SCC was 32 days (95% CI 30–42 days) in the bedaquiline susceptible cohort, 137 days (95% CI 102–172 days) earlier compared with the bedaquiline resistant cohort (Figure 1). In the univariable analysis of pooled bedaquiline-resistant cases and matched bedaquiline susceptible controls, baseline higher microscopy grade (HR 0·72, 95% CI 0·56–0·92, p=0·008), and bedaquiline resistance (HR 0·16, 95% CI 0·08–0·31, p<0·001) were associated with lower sustained SCC (Supplement Table 8). When included in a multivariable model, higher baseline microscopy (HR 0·41, 95% CI 0·23–0·73, p=0·003) and bedaquiline resistance (HR 0·06, 95% CI 0·02–0·23, p<0·001) remained significantly associated with reduced SCC. Cumulative mortality at 18 months was numerically higher at 22% (95% CI, 13%–31%) in the bedaquiline resistant group versus 18% (95% CI 9%–26%) in matched controls, but this was not statistically significant (Figure 2, log rank test p=0·5).

**Figure 2:**
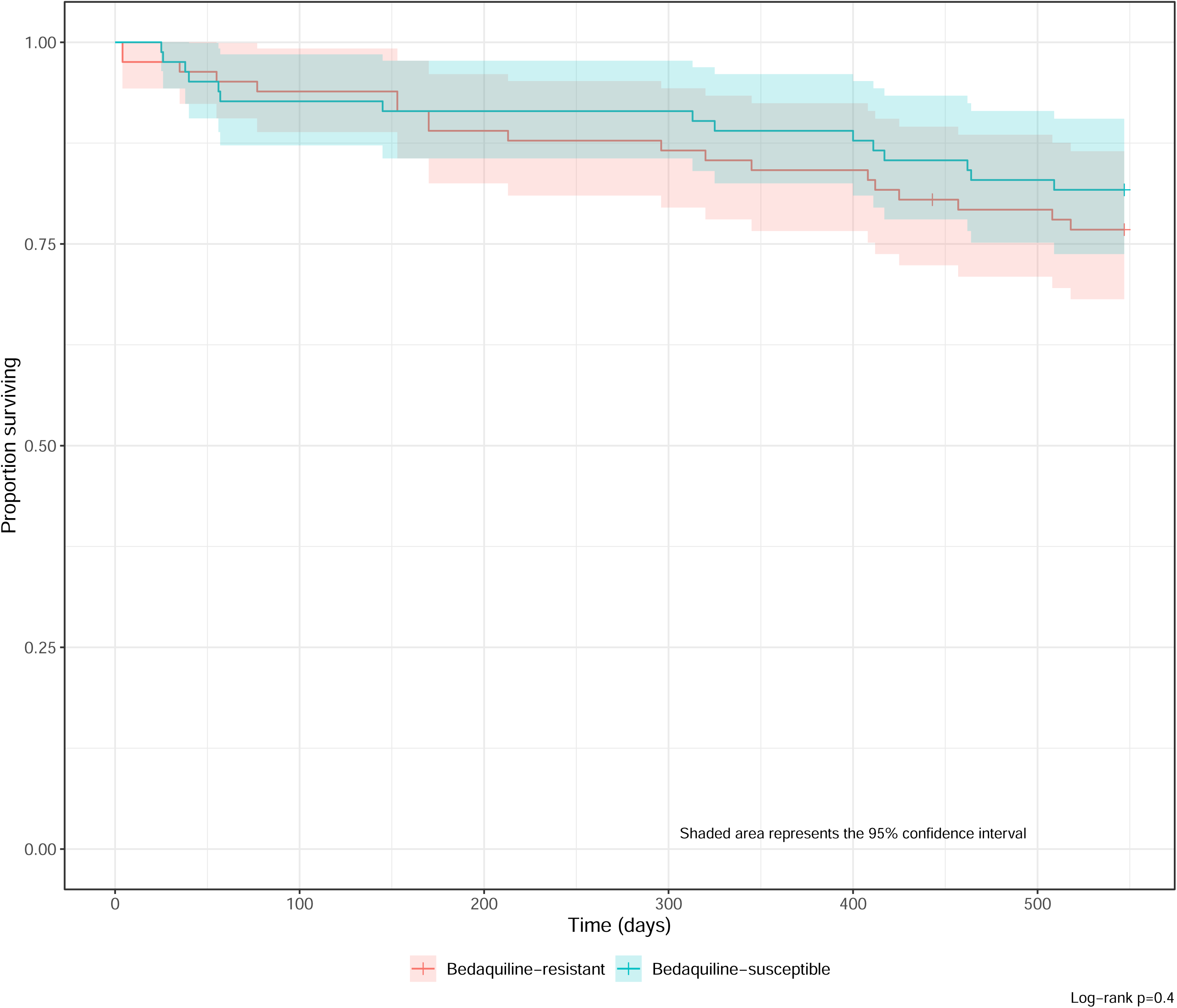
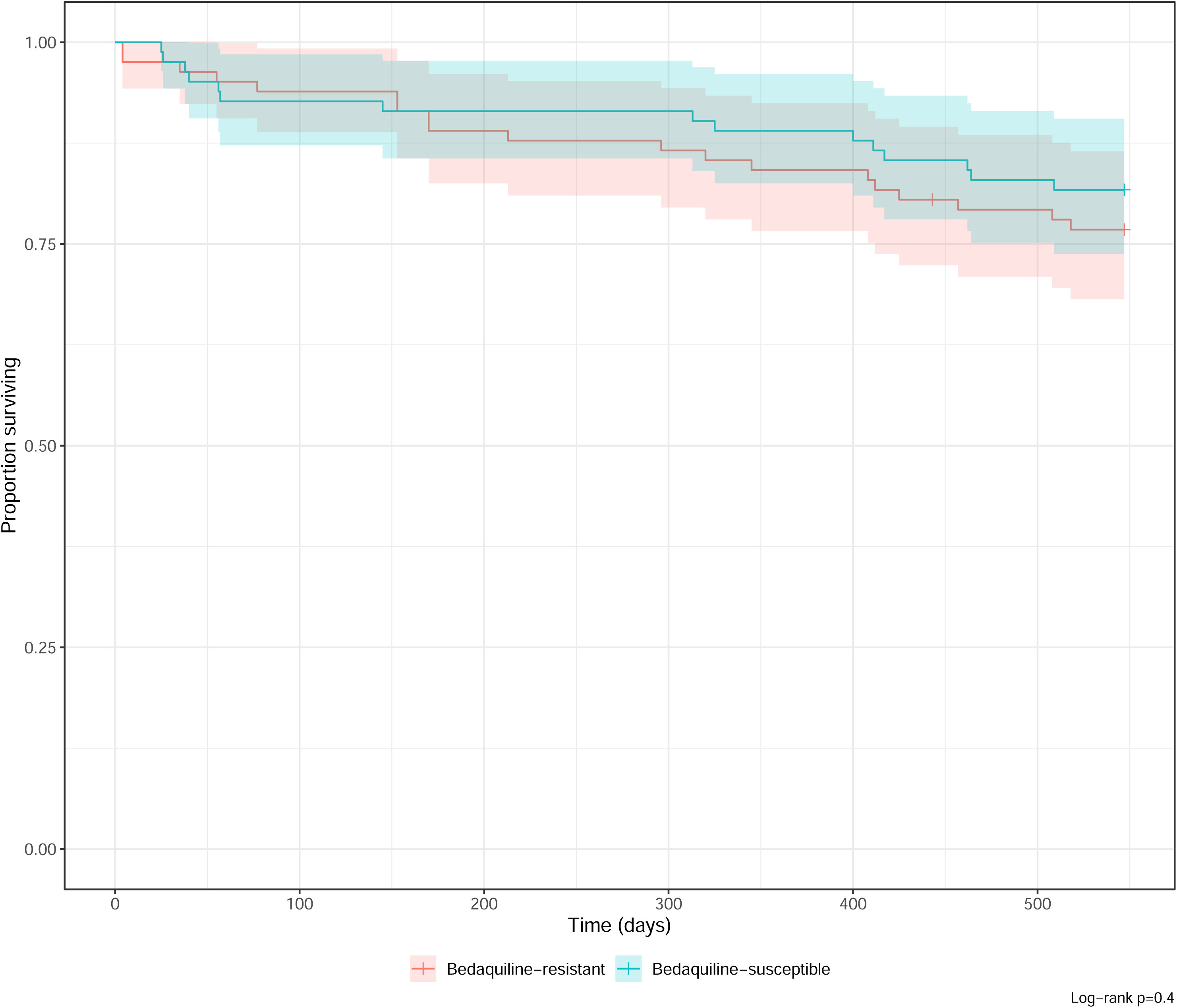
Kaplan–Meier survival curves in the combined cohort of bedaquiline–resistant cases and matched bedaquiline–susceptible controls after treatment initiation, by bedaquiline resistance and censored at 18 months.

## Discussion

Our study characterised management and long-term outcomes among patients treated for bedaquiline-resistant TB in the South African NTP. Only half of cases were alive and TB-free at 18 months after detection of bedaquiline resistance and time to SCC was almost 5 months longer than a matched RR-TB cohort without bedaquiline resistance. Treatment outcomes are comparable to those of XDR-TB (previously defined as resistance to an injectable drug and fluoroquinolones) in the pre-bedaquiline era, ^20,21^ highlighting the severe consequences of bedaquiline resistance to patients and TB programmes.

The lower microbiological treatment responses among bedaquiline resistant cases compared with matched controls observed in our study reinforces the need for an active bactericidal agent in regimens for bedaquiline-resistant TB, even when other effective Group A and Group B drugs such as linezolid and terizidone are included. Patients with bedaquiline-resistant TB in our study had much worse WHO treatment outcomes (33% favourable treatment outcome) compared with the most recent global estimates for RR-TB (68%)^1^ and those from a retrospective study in the South African NTP of unselected patients with RR-TB on bedaquiline-containing regimens (67%).^3^ In our study, this was driven by failure to culture convert and culture reversions, again highlighting the reduced effectiveness of RR-TB regimens with impaired or absent bedaquiline efficacy. An additional factor is the more extensive resistance profiles in bedaquiline-resistant isolates leading to a potentially lower overall number of effective drugs. ^10,22^ Most *M. tuberculosis* isolates from cases in our study were resistant to key second line drugs, including fluoroquinolones and clofazimine (>90% resistant). This is in contrast to a cross-sectional survey conducted across three provinces in South Africa in 2023, where 65% of bedaquiline-resistant isolates (n=149) were susceptible to fluoroquinolones. ^11^ The higher prevalence of fluoroquinolone resistance in our cohort could be an overestimate because of selection bias since bedaquiline testing was conducted based on pre-existing fluoroquinolone resistance from 2019 until March 2023 (all RR-TB cases since then are tested for bedaquiline susceptibility).

In a recent analysis of programmatic data from the South African NTP, end of treatment mortality for RR-TB with shorter bedaquiline-based regimens was around 17%, increasing over time to 24% at 24 months. ^11,23^ Similar mortality trends were seen among people with bedaquiline-resistant TB in our study. Cause of death was not ascertained, but presumably a substantial number of people experienced late mortality from uncontrolled TB disease, given that almost 20% of survivors were culture positive for *M. tuberculosis* at 18 months despite treatment. Other potential explanations for the high mortality include advanced HIV disease (over two-thirds were HIV-positive with a median CD4 count of 167 cells/mm^3^) and functional lung damage (half of cases had previous episodes of RR-TB and over a quarter experienced prior treatment failure).

Bedaquiline use is strongly associated with mortality reduction in RR-TB^24^ and resistance is therefore expected to reduce this effect. In our study cumulative mortality was numerically higher among cases with bedaquiline resistance compared with controls. Survival bias may artificially lower mortality estimates for acquired bedaquiline resistance in retrospective studies (patients may die prior to undergoing DST for bedaquiline), possibly explaining our results. Another potential explanation for attenuated mortality impact is residual treatment effect. We found an association between increasing months of bedaquiline use and reduced mortality. This observation could be partially explained by immortal time bias, but the effect was similar after excluding early deaths. Additionally, some patients had good outcomes on continued bedaquiline therapy without a carbapenem or an effective fluoroquinolone in the regimen. Bedaquiline may contribute antituberculosis effect in the context of bedaquiline resistant variants with moderate minimum inhibitory concentration (MIC) elevations, but the clinical efficacy of this strategy is unknown^25,26^ and MIC data were unavailable in our study. Fewer than half of patients were prescribed a carbapenem despite inclusion in national and WHO guideline recommendations as part of “salvage regimens” for highly resistant TB. ^6^ Carbapenem use in drug-resistant TB has been informed by observational studies that focussed on difficult to treat cases (treatment success 57%–80%),^27^ and an individual patient meta-analysis showing reduced treatment failure or relapse (adjusted odds ratio 0·4, 95% CI 0·2–0·7). ^13,28^ In our cohort, carbapenem exposure was not associated with improved outcomes, although this is limited by confounding by indication and potential allocation bias. We did not capture reasons for treatment decisions to explain relatively low carbapenem use. Plausible explanations are reluctance for intravenous therapy by care givers and patients, favourable clinical responses in some patients, and delayed recognition of bedaquiline resistance with mortality occurring before a carbapenem could be started (higher early mortality was observed among those not receiving meropenem, Supplement Figure 2).

One-fifth of cases in our study had no prior exposure to bedaquiline or clofazimine, implying new infection with bedaquiline-resistant isolates. Exogenous infection with bedaquiline resistant *M. tuberculosis* strains is well documented in South Africa^2^ with one surveillance study attributing almost 8% (n=76) of resistance cases to primary transmission. ^4^ The long delay to treatment initiation after the index sputum collection and prolonged time to SCC (median 175 days), may also contribute to community transmission, accelerating the public health threat of bedaquiline resistance. Of concern is the increasing proportion of patients in our cohort with *M. tuberculosis* isolates resistant to linezolid (from 3% to 21%), implying and additional risk of linezolid resistance amplification. Implementation of near-patient and easily interpretable rapid molecular testing, such as commercial assays for targeted next generation sequencing, for detecting bedaquiline resistance (and for companion drugs) is a priority for TB programmes to support management and reduce risk time for transmission of bedaquiline resistance.

Our study had limitations. Patients were identified from a single treatment centre, impacting generalisability because of differences in clinical management, host factors (e.g. HIV prevalence), and infecting *M. tuberculosis* strains. The retrospective study design resulted in missing data and impacted on quality of outcome information; however, we captured outcomes reported by the NTP, thereby reflecting routine care. Although we matched on important prognostic factors, the comparison is imperfect because of residual differences in the populations that may confound outcomes (e.g. no prior RR-TB in the matched controls). Despite this, the analysis provides some insight on the independent effect of bedaquiline resistance. Timing of treatment initiation directed towards bedaquiline-resistant TB was unreliably documented in medical records. We therefore selected time of index sputum collection for analysis of the key clinical outcomes to reflect real-world experience and avoid survival bias associated with delays in treatment start. Finally, during the study period, bedaquiline resistance testing was mainly offered to patients with poor treatment response or fluoroquinolone resistance. This may bias towards a sicker population, overestimating the impact of bedaquiline resistance on TB treatment outcomes. The South African NTP now recommends routine bedaquiline DST for all RR-TB cases and continued surveillance is necessary to determine more representative outcomes. ^6^

In conclusion, our study shows that people with bedaquiline-resistant TB have extremely limited treatment options and, consequently, suffer poor treatment outcomes despite prolonged antituberculosis therapy with available drugs and good retention in care. As bedaquiline resistance expands, there is an urgent need to introduce diagnostics for early and near-patient bedaquiline resistance detection and to identify evidence-based treatment options with new drug classes, to avoid a return to a pre-bedaquiline era.

## Data Availability

Individual deidentified patient data from the bedaquiline resistant cohort in the present study including a data dictionary will be available upon request after contact with the corresponding author at publication after approval of study proposal and a signed data sharing agreement.

## Acknowledgement

We thank and acknowledge Tracy Glass from the South African Medical Research Council who assisted with linkage to the South African Population Register for people with unknown vital status at follow-up.

## Data sharing

Individual deidentified patient data from the bedaquiline resistant cohort including a data dictionary will be available upon request after contact with the corresponding author at publication after approval of study proposal and a signed data sharing agreement.

## Contributions

SW, NM, and GM conceptualized the study with methodology input from RW, JK, ZM, JAMS and LM. LM and ZM collected the data with data curation conducted by JK. RW conducted microbiological analyses. JK and SW supervised the study. ZM performed the analysis and visualisation. LM, ZM, JAMS, SW, and JK had full access to the data while ZM and JK have directly verified the underlying data. JK, LM, ZM and SW wrote the original draft; all coauthors reviewed and edited the manuscript. All authors approved the final draft of the manuscript.

## References

1. World Health Organization. Global Tuberculosis Report 2024. 2024. https://www.who.int/teams/global-tuberculosis-programme/tb-reports/global-tuberculosis-report-2024 (accessed 19 December 2024).

2. Derendinger B, Dippenaar A, de Vos M, et al. Bedaquiline resistance in patients with drug-resistant tuberculosis in Cape Town, South Africa: a retrospective longitudinal cohort study. Lancet Microbe 2023; 4(12): e972–e82.

3. Pai H, Ndjeka N, Mbuagbaw L, et al. Bedaquiline safety, efficacy, utilization and emergence of resistance following treatment of multidrug-resistant tuberculosis patients in South Africa: a retrospective cohort analysis. BMC Infect Dis 2022; 22(1): 870.

4. Ismail NA, Omar SV, Moultrie H, et al. Assessment of epidemiological and genetic characteristics and clinical outcomes of resistance to bedaquiline in patients treated for rifampicin-resistant tuberculosis: a cross-sectional and longitudinal study. Lancet Infect Dis 2022; 22(4): 496–506.

5. Department of Health: Republic of South Africa. Management of rifampicin-resistant tuberculosis: A clinical reference guide. November 2019. https://www.health.gov.za/wp-content/uploads/2020/11/management-of-rifampicin-resistant-tb-booklet-0220-v11.pdf (accessed 15 November 2024).

6. Department of Health: Republic of South Africa. Clinical management of rifampicin- resistant Tuberculosis: Updated clinical reference guide. September 2023. https://www.health.gov.za/wp-content/uploads/2023/10/Updated-RR-TB-Clinical-Guidelines-September-2023.pdf (accessed 15 November 2024).

7. Villellas C, Stevenaert F, Remmerie B, Andries K. Sub-MIC levels of bedaquiline and clofazimine can select Mycobacterium tuberculosis mutants with increased MIC. Antimicrob Agents Chemother 2024; 68(4): e0127523.

8. de Vos M, Ley SD, Wiggins KB, et al. Bedaquiline Microheteroresistance after Cessation of Tuberculosis Treatment. N Engl J Med 2019; 380(22): 2178–80.

9. Nimmo C, Millard J, van Dorp L, et al. Population-level emergence of bedaquiline and clofazimine resistance-associated variants among patients with drug-resistant tuberculosis in southern Africa: a phenotypic and phylogenetic analysis. Lancet Microbe 2020; 1(4): e165–e74.

10. Timm J, Bateson A, Solanki P, et al. Baseline and acquired resistance to bedaquiline, linezolid and pretomanid, and impact on treatment outcomes in four tuberculosis clinical trials containing pretomanid. PLOS Glob Public Health 2023; 3(10): e0002283.

11. Moultrie H, Kachingwe E, Ismail F, Vally Omar S. Bedaquiline susceptibility surveillance using routine laboratory data, South Africa (July 2019-November 2023). 2024.

12. Barilar I, Fernando T, Utpatel C, et al. Emergence of bedaquiline-resistant tuberculosis and of multidrug-resistant and extensively drug-resistant Mycobacterium tuberculosis strains with rpoB Ile491Phe mutation not detected by Xpert MTB/RIF in Mozambique: a retrospective observational study. Lancet Infect Dis 2024; 24(3): 297–307.

13. World Health Organization. WHO consolidated guidelines on tuberculosis. Module 4: treatment - drug-resistant tuberculosis treatment, 2022 update. 2022. https://www.who.int/publications/i/item/9789240007048 (accessed 10 November 2024).

14. World Health Organization. Meeting report of the WHO expert consultation on the definition of extensively drug-resistant tuberculosis, 27-29 October 2020. 2021. https://www.who.int/publications/i/item/9789240018662 (accessed 14 November 2024).

15. Seung KJ, Becerra MC, Atwood SS, Alcantara F, Bonilla CA, Mitnick CD. Salvage therapy for multidrug-resistant tuberculosis. Clin Microbiol Infect 2014; 20(5): 441–6.

16. Stadler J, Wasserman S, Meintjes G, Warren R. Treatment outcomes with a shorter all-oral regimen for rifampicin-resistant TB in a programmatic setting in South Africa: interim analysis from the SHIFT-TB cohort. World Conference on Lung Health; 2023 15-18 November; Paris: The International Journal of Tuberculosis and Lung Disease (IJTLD); 2023. p. S1-S725.

17. Shapiro AN, Scott L, Moultrie H, et al. Tuberculosis testing patterns in South Africa to identify groups that would benefit from increased investigation. Sci Rep 2023; 13(1): 20875.

18. National Health Laboratory Service (NHLS) [of South Africa]. Standard Operating Procedure: MGIT 960 DST procedure. Johannesburg: NHLS, 2023.

19. Kurbatova EV, Taylor A, Gammino VM, et al. Predictors of poor outcomes among patients treated for multidrug-resistant tuberculosis at DOTS-plus projects. Tuberculosis (Edinb) 2012; 92(5): 397–403.

20. Shah NS, Pratt R, Armstrong L, Robison V, Castro KG, Cegielski JP. Extensively drug-resistant tuberculosis in the United States, 1993-2007. JAMA 2008; 300(18): 2153–60.

21. Pietersen E, Ignatius E, Streicher EM, et al. Long-term outcomes of patients with extensively drug-resistant tuberculosis in South Africa: a cohort study. Lancet 2014; 383(9924): 1230–9.

22. CRyPTIC Consortium. Epidemiological cut-off values for a 96-well broth microdilution plate for high-throughput research antibiotic susceptibility testing of M. tuberculosis. Eur Respir J 2022; 60(4).

23. Ndjeka N, Campbell JR, Meintjes G, et al. Treatment outcomes 24 months after initiating short, all-oral bedaquiline-containing or injectable-containing rifampicin-resistant tuberculosis treatment regimens in South Africa: a retrospective cohort study. Lancet Infect Dis 2022; 22(7): 1042–51.

24. Schnippel K, Ndjeka N, Maartens G, et al. Effect of bedaquiline on mortality in South African patients with drug-resistant tuberculosis: a retrospective cohort study. Lancet Respir Med 2018; 6(9): 699–706.

25. Pym AS, Diacon AH, Tang SJ, et al. Bedaquiline in the treatment of multidrug- and extensively drug-resistant tuberculosis. Eur Respir J 2016; 47(2): 564–74.

26. Nguyen TVA, Anthony RM, Banuls AL, Nguyen TVA, Vu DH, Alffenaar JC. Bedaquiline Resistance: Its Emergence, Mechanism, and Prevention. Clin Infect Dis 2018; 66(10): 1625–30.

27. Sotgiu G, D’Ambrosio L, Centis R, et al. Carbapenems to Treat Multidrug and Extensively Drug-Resistant Tuberculosis: A Systematic Review. Int J Mol Sci 2016; 17(3): 373.

28. Collaborative Group for the Meta-Analysis of Individual Patient Data, Ahmad N, Ahuja SD, et al. Treatment correlates of successful outcomes in pulmonary multidrug-resistant tuberculosis: an individual patient data meta-analysis. Lancet 2018; 392(10150): 821–34.

