## Supplement for "Treatment outcomes of bedaquiline-resistant tuberculosis: a retrospective and matched cohort study"

Supplementary Methods	3

Supplement Table 1: Drug susceptibility testing of sputum and proportion with resistance to each drug before, at, and after the index sputum^1^ collection when bedaquiline resistance was identified	4

Supplement Table 2: Univariable analysis of predictors of time to sustained sputum culture conversion at 12 months after treatment initiation following index sputum^1^ collection in patients with bedaquiline-resistant tuberculosis using a Cox proportional hazards model	5

Supplement Table 3: Tuberculosis-free survival at 6, 12, and 18 months after treatment initiation following the index sputum^1^ collection in patients with bedaquiline-resistant tuberculosis	6

Supplement Table 4: Univariable and multivariable analysis of predictors of time to death over 18 months from index sputum^1^ collection in patients with bedaquiline-resistant tuberculosis using a Cox proportional hazards model	7

Supplement Table 5: Univariable and multivariable analysis of predictors of time to death over 18 months from index sputum^1^ collection in patients with bedaquiline-resistant tuberculosis who survived at least eight weeks after the index sputum^1^ collection date using a Cox proportional hazards model (sensitivity analysis)	8

Supplement Table 6: Modified World Health Organization treatment outcomes after treatment initiation of following the index sputum^1^ collection in patients with bedaquiline-resistant tuberculosis	9

Supplement Table 7: Participant and clinical characteristics of the matched controls with bedaquiline-susceptible rifampicin-resistant tuberculosis at the time of treatment initiation	10

Supplement Table 8: Univariable and multivariable analysis of time to sustained sputum culture conversion after treatment initiation in the combined cohort of bedaquiline-resistant cases and matched bedaquiline-susceptible controls using a stratified Cox proportional hazard model	11

Supplement Figure 1: Kaplan-Meier curves for time to first sputum culture conversion from treatment initiation in the combined cohort of bedaquiline-resistant cases and matched bedaquiline-susceptible controls, by bedaquiline resistance and censored at 12 months	12

Supplement Figure 2: Kaplan-Meier curves of survival from time of index sputum^1^ collection for patients with bedaquiline resistant tuberculosis, by carbapenem use and censored at 18 months.	13

References	14

### Supplementary Methods

Unfavourable World Health Organization (WHO) treatment outcome was adapted from the WHO 2021 definition^1^ with the following differences: 1) treatment failure due lack of sputum culture conversion at six months or reversion at any timepoint was assigned regardless of whether the treatment regimen was changed; and 2) for patients whose regimens were changed due to any baseline drug resistance, outcomes were reported from the time of regimen change, not from treatment start.

The control group with confirmed phenotypic bedaquiline susceptibility was matched to bedaquiline-resistant cases based on age, HIV status, and baseline culture status. Age matching was conducted by categorising patients into the following age groups: 10–20 years, 20–30 years, 30–40 years, 40–50 years, 50–60 years, and over 60 years.

### Supplement Table 1: Drug susceptibility testing of sputum and proportion with resistance to each drug before, at, and after the index sputum^1^ collection when bedaquiline resistance was identified

| **Characteristic** | **Before the index sputum***^1^*  N = 45*^2^* | **At the index sputum***^1^*  N = 82*^2^* | **After the index sputum***^1^*  N = 45*^2^* |
| --- | --- | --- | --- |
| **Xpert MTB/Rif Ultra** |  |  |  |
| Rifampicin | 9 (90%), n = 10 | 18 (95%), n = 19 | 8 (100%), n = 8 |
| **Line Probe Assay** |  |  |  |
| Rifampicin | 44 (98%), n = 45 | 72 (97%), n = 74 | 42 (100%), n = 42 |
| Isoniazid | 44 (98%), n = 45 | 70 (95%), n = 74 | 42 (98%), n = 43 |
| Fluoroquinolone | 28 (72%), n = 39 | 61 (88%), n = 69 | 37 (86%), n = 43 |
| **Phenotypic DST** |  |  |  |
| Amikacin | 5 (63%), n = 8 | 9 (53%), n = 17 | 7 (70%), n = 10 |
| Bedaquiline | 1 (5·0%), n = 20 | 82 (100%), n = 82 | 28 (93%), n = 30 |
| Clofazimine | 2 (18%), n = 11 | 67 (92%), n = 73 | 19 (90%), n = 21 |
| Delaminid | 0, n = 0 | 1 (100%), n = 1 | 0, n = 0 |
| Ethambutol | 1 (100%), n = 1 | 5 (71%), n = 7 | 4 (80%), n = 5 |
| Ethionamide | 0 (0%), n = 2 | 9 (82%), n = 11 | 6 (86%), n = 7 |
| Isoniazid | 3 (100%), n = 3 | 10 (91%), n = 11 | 5 (83%), n = 6 |
| Isoniazid, high dose | 2 (67%), n = 3 | 8 (100%), n = 8 | 5 (83%), n = 6 |
| Levofloxacin | 1 (17%), n = 6 | 15 (68%), n = 22 | 7 (64%), n = 11 |
| Linezolid | 2 (7·1%), n = 28 | 2 (2·6%), n = 77 | 7 (21%), n = 33 |
| Moxifloxacin | 12 (86%), n = 14 | 24 (92%), n = 26 | 8 (80%), n = 10 |
| Moxifloxacin, high dose | 12 (80%), n = 15 | 21 (66%), n = 32 | 7 (54%), n = 13 |
| Para-amino salicylic acid | 0 (0%), n = 2 | 0 (0%), n = 12 | 2 (22%), n = 9 |
| Pyrazinamide | 1 (33%), n = 3 | 4 (80%), n = 5 | 5 (83%), n = 6 |
| Rifabutin | 1 (50%), n = 2 | 9 (69%), n = 13 | 8 (80%), n = 10 |
| *^1^*Index sputum is defined as the first bedaquiline-resistant *Mycobacterium tuberculosis* isolate  *^2^*n (%), n = N, DST = Drug susceptibility testing | | | |

### Supplement Table 2: Univariable analysis of predictors of time to sustained sputum culture conversion at 12 months after treatment initiation following index sputum^1^ collection in patients with bedaquiline-resistant tuberculosis using a Cox proportional hazards model

| **Characteristic** | **Number**  **of events** | **HR** | **95% CI** | **p-value** |
| --- | --- | --- | --- | --- |
| Age | 50 | 1.01 | 0.99, 1.03 | 0.26 |
| Sex |  |  |  |  |
| Male | 24 | ·· | ·· |  |
| Female | 26 | 1.02 | 0.59, 1.78 | 0.93 |
| BMI | 49 | 1.04 | 0.98, 1.10 | 0.22 |
| HIV |  |  |  |  |
| No | 13 | ·· | ·· |  |
| Yes | 37 | 1.43 | 0.76, 2.69 | 0.27 |
| Combined HIV and ART status |  |  |  |  |
| HIV negative | 13 | ·· | ·· |  |
| HIV positive, on ART | 34 | 1.48 | 0.78, 2.80 | 0.23 |
| HIV positive, not on ART | 3 | 1.05 | 0.30, 3.67 | 0.94 |
| Previous RR-TB |  |  |  |  |
| No | 24 | ·· | ·· |  |
| Yes | 26 | 1.03 | 0.59, 1.80 | 0.90 |
| Baseline microscopy status |  |  |  |  |
| Negative | 25 | ·· | ·· |  |
| Positive | 25 | 0.95 | 0.54, 1.66 | 0.86 |
| Bedaquiline use in the index episode |  |  |  |  |
| No | 4 | ·· | ·· |  |
| Yes | 46 | 2.02 | 0.73, 5.63 | 0.18 |
| ^1^Index sputum is defined as the first bedaquiline-resistant Mycobacterium tuberculosis isolate  ART = Antiretroviral treatment, CI = Confidence Interval, HR = Hazard Ratio, RR-TB = Rifampicin-resistant tuberculosis | | | | |

### Supplement Table 3: Tuberculosis-free survival at 6, 12, and 18 months after treatment initiation following the index sputum^1^ collection in patients with bedaquiline-resistant tuberculosis

| **Characteristic** | **Month 6** N = 82*^2^* | **Month 12** N = 82*^2^* | **Month 18** N = 81*^2, 3^* |
| --- | --- | --- | --- |
| Achieved | 45 (55%) | 47 (57%) | 41 (51%) |
| Alive, TB free & treatment complete*^4^* | 0 (0%) | 2 (2.4%) | 8 (9.8%) |
| Alive, TB free & in care (treatment ongoing) | 45 (55%) | 45 (55%) | 33 (41%) |
| Not achieved | 37 (45%) | 35 (43%) | 40 (49%) |
| Not alive (i.e. died) | 8 (9.8%) | 13 (16%) | 19 (23%) |
| Not TB free, alive & in care (treatment ongoing) | 23 (28%) | 18 (22%) | 16 (20%) |
| Not in care*^5^* | 6 (7.3%) | 4 (4.9%) | 5 (6.1%) |
| Alive, TB free | 2 (2.4%) | 2 (2.4%) | 3 (3.7%) |
| Alive, not TB free | 4 (4.9%) | 2 (2.4%) | 2 (2.4%) |
| *^1^*Index sputum is defined as the first bedaquiline-resistant Mycobacterium tuberculosis isolate  ^2^n (%)  *^3^*18 months post-treatment initiation had not yet passed for one participant at the date of database closure  ^4^TB-free is defined as achieving sputum culture conversion without culture reversion by the specified timepoint.  ^5^Lost to follow up (irrespective of TB status) or completed treatment but not TB-free.  TB = Tuberculosis | | | |

### Supplement Table 4: Univariable and multivariable analysis of predictors of time to death over 18 months from index sputum^1^ collection in patients with bedaquiline-resistant tuberculosis using a Cox proportional hazards model

|  | **Univariable Analysis** | | | |
| --- | --- | --- | --- | --- |
| **Characteristic** | **Event N** | **HR** | **95% CI** | **p-value** |
| Age | 19 | 1.01 | 0.97, 1.05 | 0.64 |
| Sex |  |  |  |  |
| Male | 10 | — | — |  |
| Female | 9 | 0.83 | 0.34, 2.05 | 0.69 |
| BMI | 19 | 0.88 | 0.77, 1.01 | 0.070 |
| HIV |  |  |  |  |
| No | 6 | — | — |  |
| Yes | 13 | 0.94 | 0.36, 2.46 | 0.89 |
| Combined HIV and ART |  |  |  |  |
| HIV negative | 6 | — | — |  |
| HIV positive, on ART | 11 | 0.88 | 0.32, 2.38 | 0.80 |
| HIV positive, not on ART | 2 | 1.45 | 0.29, 7.20 | 0.65 |
| Baseline microscopy |  |  |  |  |
| Negative | 7 | — | — |  |
| Positive | 12 | 1.57 | 0.62, 3.99 | 0.34 |
| Previous RR-TB |  |  |  |  |
| No | 10 | — | — |  |
| Yes | 9 | 0.90 | 0.36, 2.21 | 0.81 |
| CD4 count |  |  |  |  |
| < 200 | 5 | — | — |  |
| ≥ 200 | 4 | 0.91 | 0.24, 3.37 | 0.88 |
| Unknown | 4 | 0.95 | 0.25, 3.54 | 0.94 |
| Duration of bedaquiline (months) | 19 | 0.75 | 0.63, 0.90 | 0.002 |
| *^1^*Index sputum is defined as the first bedaquiline-resistant *Mycobacterium tuberculosis* isolate  HR = Hazard Ratio, CI = Confidence Interval | | | | |

### Supplement Table 5: Univariable and multivariable analysis of predictors of time to death over 18 months from index sputum^1^ collection in patients with bedaquiline-resistant tuberculosis who survived at least eight weeks after the index sputum^1^ collection date using a Cox proportional hazards model (sensitivity analysis)

|  | **Univariable Analysis** | | | | **Multivariable Analysis** | | |
| --- | --- | --- | --- | --- | --- | --- | --- |
| **Characteristic** | **Event N** | **HR** | **95% CI** | **p-value** | **HR** | **95% CI** | **p-value** |
| Age | 15 | 1.02 | 0.98, 1.06 | 0.33 |  |  |  |
| Sex |  |  |  |  |  |  |  |
| Male | 7 | — | — |  |  |  |  |
| Female | 8 | 1.05 | 0.38, 2.90 | 0.92 |  |  |  |
| BMI | 15 | 0.84 | 0.71, 0.99 | 0.040 | 0.86 | 0.80, 1.01 | 0.071 |
| HIV |  |  |  |  |  |  |  |
| No | 5 | — | — |  |  |  |  |
| Yes | 10 | 0.85 | 0.29, 2.50 | 0.77 |  |  |  |
| Combined HIV and ART |  |  |  |  |  |  |  |
| HIV negative | 5 | — | — |  |  |  |  |
| HIV positive, on ART | 8 | 0.76 | 0.25, 2.31 | 0.62 |  |  |  |
| HIV positive, not on ART | 2 | 1.77 | 0.34, 9.10 | 0.50 |  |  |  |
| Baseline microscopy |  |  |  |  |  |  |  |
| Negative | 6 | — | — |  |  |  |  |
| Positive | 9 | 1.39 | 0.49, 3.90 | 0.54 |  |  |  |
| Previous RR-TB |  |  |  |  |  |  |  |
| No | 9 | — | — |  |  |  |  |
| Yes | 6 | 0.66 | 0.23, 1.85 | 0.43 |  |  |  |
| CD4 count |  |  |  |  |  |  |  |
| < 200 | 3 | — | — |  |  |  |  |
| ≥ 200 | 4 | 1.49 | 0.33, 6.64 | 0.60 |  |  |  |
| Unknown | 3 | 1.15 | 0.23, 5.72 | 0.86 |  |  |  |
| Duration of bedaquiline (months) | 15 | 0.82 | 0.69, 0.99 | 0.039 | 0.85 | 0.71, 1.01 | 0.069 |
| ^1^Index sputum is defined as the first bedaquiline-resistant *Mycobacterium tuberculosis* isolate  ART = Antiretroviral treatment, CI = Confidence Interval, HR = Hazard Ratio, RR-TB = Rifampicin-resistant tuberculosis | | | | | | | |

### Supplement Table 6: Modified World Health Organization treatment outcomes after treatment initiation of following the index sputum^1^ collection in patients with bedaquiline-resistant tuberculosis

| **World Health Organization Treatment Outcome** | **N = 81** *^2,3^* |
| --- | --- |
| Favourable treatment outcome | 27 (33%) |
| Cured | 24 (30%) |
| Treatment completed | 3 (3·7%) |
| Unfavourable treatment outcome | 54 (67%) |
| Died | 11 (14%) |
| Lost to follow-up | 8 (9·9%) |
| Treatment failed | 35 (43%) |
| Reason for treatment failure |  |
| Lack of culture conversion by month 6 | 14 (40%) |
| Culture reversion | 10 (29%) |
| Change of regimen and/or permanently changed ≥2 drugs | 11 (31%) |
| Reason for regimen change |  |
| Poor clinical response and/or no bacteriological response based on clinical judgement | 11 (100%) |
| ^1^Index sputum is defined as the first bedaquiline-resistant *Mycobacterium tuberculosis* isolate  *^2^*n (%)  ^3^One patient was not yet assigned a World Health Organization outcome due to ongoing treatment at the end of the study (20 January 2025) and was excluded from this analysis. | |

### Supplement Table 7: Participant and clinical characteristics of the matched controls with bedaquiline-susceptible rifampicin-resistant tuberculosis at the time of treatment initiation

| **Characteristic** | **N = 82***^1^* |
| --- | --- |
| Age, years | 37 (32, 44) (18, 61) |
| Female sex | 37 (45%) |
| Weight, kg | 52 (46, 60) (35, 97) |
| BMI, kg/m2 | 18.2 (16.6, 21.5) (12.0, 38.4) |
| Sputum microscopy positive | 59 (72%) |
| Baseline microscopy grade |  |
| 0 | 23 (28%) |
| 1+ | 18 (22%) |
| 2+ | 20 (24%) |
| 3+ | 21 (26%) |
| HIV positive | 57 (70%) |
| ART status (if HIV positive) |  |
| Currently on ART | 32 (56%) |
| Not on ART | 25 (44%) |
| HIV viral load^2^ (if HIV positive) |  |
| Below level of detection | 13 (25%) |
| Detectable | 38 (75%) |
| Quantitative viral load^2^, n=42 | 42,750 (200, 592500) (0, 2,538600) |
| CD4 count^2^, cells/mm³, n=49 | 100 (27, 210) (2, 682) |
| Number of medications at treatment initiation | 7 (7, 7) (5, 7) |
| Confirmed susceptiblity to bedaquiline | 3 (9·4%) |
| *^1^*Median (Q1, Q3) (Min, Max); n (%)  *^2^*Within six months before and after treatment initiation  ART = Antiretroviral treatment, BMI = Body mass index | |

### Supplement Table 8: Univariable and multivariable analysis of time to sustained sputum culture conversion after treatment initiation in the combined cohort of bedaquiline-resistant cases and matched bedaquiline-susceptible controls using a stratified Cox proportional hazard model

|  | **Univariate Analysis** | | | | **Multivariate Analysis** | | |
| --- | --- | --- | --- | --- | --- | --- | --- |
| **Characteristic** | **Event N** | **HR** | **95% CI** | **p-value** | **HR** | **95% CI** | **p-value** |
| Sex |  |  |  |  |  |  |  |
| Male | 63 | ·· | ·· |  |  |  |  |
| Female | 59 | 1.07 | 0.52, 2.22 | 0.85 |  |  |  |
| BMI | 121 | 0.98 | 0.91, 1.06 | 0.62 |  |  |  |
| Baseline microscopy grade | 122 | 0.72 | 0.56, 0.92 | 0.008 | 0.41 | 0.23, 0.73 | 0.003 |
| Baseline fluoroquinolone resistance |  |  |  |  |  |  |  |
| No | 9 | ·· | ·· |  |  |  |  |
| Yes | 94 | 1.00 | 0.29, 3.45 | >0.99 |  |  |  |
| Bedaquiline resistance |  |  |  |  |  |  |  |
| No | 72 | ·· | ·· |  | ·· | ·· |  |
| Yes | 50 | 0.16 | 0.08, 0.31 | <0.001 | 0.06 | 0.02, 0.23 | <0.001 |
| HR = Hazard Ratio, CI = Confidence Interval | | | | | | | |

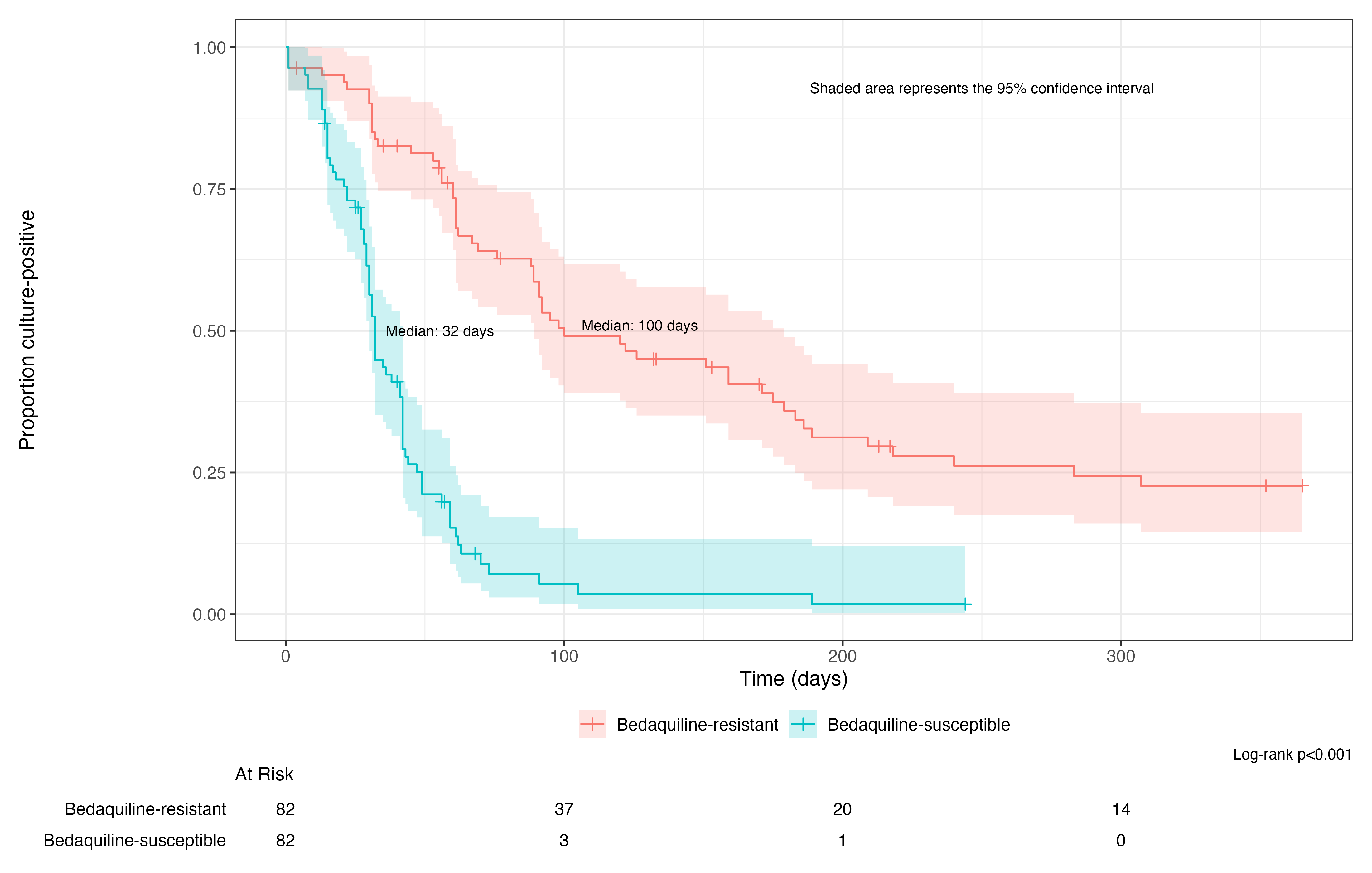

### Supplement Figure 1: Kaplan-Meier curves for time to first sputum culture conversion from treatment initiation in the combined cohort of bedaquiline-resistant cases and matched bedaquiline-susceptible controls, by bedaquiline resistance and censored at 12 months

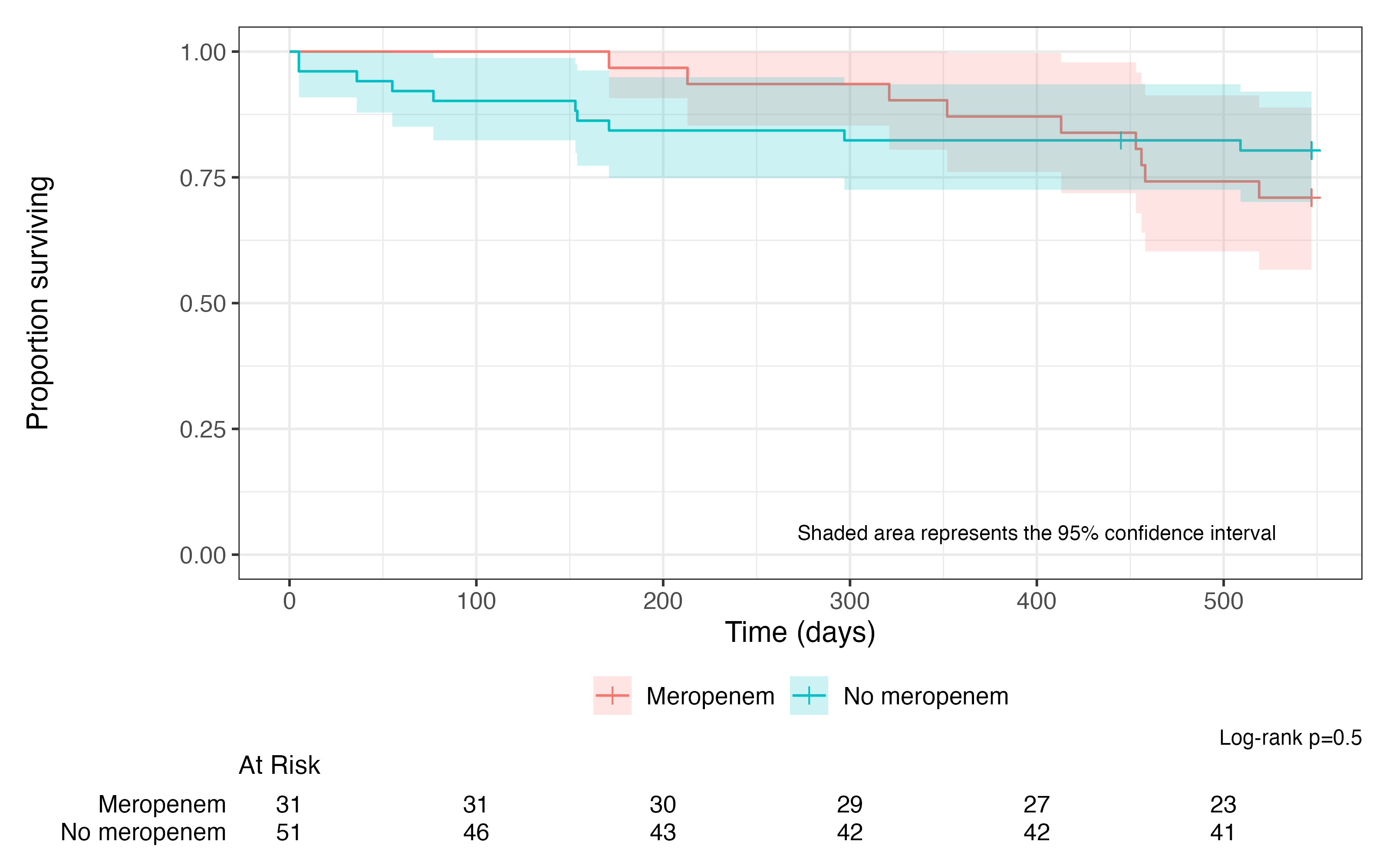

### Supplement Figure 2: Kaplan-Meier curves of survival from time of index sputum^1^ collection for patients with bedaquiline resistant tuberculosis, by carbapenem use and censored at 18 months.

*^1^*Index sputum is defined as the first bedaquiline-resistant *Mycobacterium tuberculosis* isolate

### References

1.	World Health Organization. WHO operational handbook on tuberculosis. Module 4: Treatment - drug-resistant tuberculosis treatment, 2022 update. Geneva: WHO, 2022.
